# Historical Perspectives in Medicine using a Large Language Model: Emulating an 18th Century Physician

**DOI:** 10.64898/2026.02.10.26345990

**Authors:** Pranav Malladi, Kasie Roark, Jennifer Eaton, Ezequiel Gleichgerrcht, Ioulia Chatzistamou, S.Wright Kennedy, Leonardo Bonilha

## Abstract

**Introduction:** Eighteenth-century medical texts document a formative period in the evolution of clinical reasoning, yet their integration into modern medical education is limited. The traditional approach to learning the history of medicine has naturally focused on passive reading, but new approaches using AI could enable learners to interrogate and simulate the historical diagnostic logic and therapeutic paradigms. More specifically, large language models (LLMs) offer an opportunity to create interactive simulations that allow experiential engagement with historical medical reasoning.

**Methods:** We developed a historically constrained LLM-based educational platform designed to emulate the diagnostic reasoning, language, and conceptual frameworks of an 18th-century physician. A modern GPT architecture was customized using strict instruction-based constraints and limited exclusively to a curated corpus of six foundational 17th–18th century medical texts. Guardrails were implemented to prevent anachronistic terminology and modern medical concepts. Model outputs were evaluated qualitatively by comparing the model’s diagnoses and treatment plans with published diagnoses and treatment from original 18^th^ century sources. We also applied the simulation to modern clinical vignettes for an illustrative contrast between modern and 18^th^ century approaches.

**Results:** The model generated responses that closely aligned with 18th-century medical and rhetorical style, as well as therapeutic reasoning. When presented with historical cases, the simulation demonstrated strong concordance with original diagnoses and management strategies. Secondly, when applied to modern cases, the model described period-appropriate reasoning, highlighting clear contrasts with contemporary biomedical reasoning.

**Conclusions:** AI broadly, and more specifically LLMs configured as historically constrained simulators, can function as effective tools for learning in medical history. This approach could enable active engagement with historical clinical reasoning, fostering critical reflection on the contingent and evolving nature of medical knowledge. Such temporal simulations hold promise for medical humanities education and interdisciplinary teaching.

## Introduction

The 18th century marked a turning point in the history of medicine, characterized by a shift from theoretical inference to a greater emphasis on observation and experimental practices[1]. The 18th century also saw a significant increase in the number of medical textbooks. The printing press, which had been in use since the 15th century, became more efficient, leading to a greater availability and distribution of medical texts across Europe and the Americas[2]. Many of these documents have survived to this day, and they not only reflect the medical knowledge available to 18th-century physicians but also the cultural, social, and intellectual contexts in which they operated[3].

Yet, many medical discoveries that shape modern medicine were not identified until the 19th century, such as the invention of the stethoscope (René Laennec in 1816), the isolation of morphine (Friedrich Sertürner in 1805) and anesthesia (William T.G. Morton in 1846), handwashing (Ignaz Semmelweis in 1847), germ theory of disease (Louis Pasteur in the 1850s and 1860s), antiseptic surgery (Joseph Lister in 1865), hypodermic syringe (Charles Pravaz in 1853), bacteriology (Robert Koch in the 1870s and 1880s), rabies vaccine (Louis Pasteur in 1885), X-rays (Wilhelm Röntgen in 1895), and aspirin (Felix Hoffmann’s in 1897), to name a few.

Thus, the 18th century represents a transitional period in which medical knowledge became increasingly accessible through an expanding corpus of texts, yet remained rooted in pre-scientific frameworks and humoral explanatory models[1, 2, 4, 5].

In recent years, advances in artificial intelligence (AI) and large language models (LLMs) have opened new avenues for exploring historical texts and simulating human-like interactions. While LLMs are trained primarily on contemporary texts and reflect modern language structures, they can be fine-tuned or adapted to align with historical linguistic patterns. Their ability to generate grammatically coherent sentences provides a solid foundation for reconstructing different stylistic and rhetorical conventions of different time periods.

Moreover, despite being trained on modern datasets, LLMs can be constrained to provide responses based solely on specific historical texts, ensuring that their outputs remain faithful to the knowledge and perspectives of a given era. In cases where the requested information is absent from the source material, these models can be designed to respond with an appropriate “I don’t know,” reinforcing the authenticity of their historical representation.

In a 1985 speech at Svaneholm Castle, Sweden, Steve Jobs famously shared his vision for leveraging technology to immortalize the intellect of great minds. He stated his hope that future generations could “ask Aristotle a question and get an answer”[6]. Our work explores an early pedagogical realization of this idea within a medical context.

Leveraging new innovations in AI, we aimed to create a platform where the user can experience an interaction with an 18th-century physician. More specifically, we created an AI-driven historical simulation adapted using instruction-based constraints related exclusively to 18th-century medical documents, enabling us to simulate the communication and decision-making processes of a physician from that time. Through this approach, learners can explore how 18th-century physicians may have approached diagnosis and treatment using period-appropriate language, offering a unique and interactive perspective on historical medical practice.

Beyond their utility for contemporary medical education, LLMs present a unique opportunity to function as temporal simulators, i.e., interactive tools that allow learners to engage dynamically with past published clinical reasoning frameworks. Traditional historical texts provide static descriptions of how 18^th^-century physicians thought. Conversely, LLMs configured as historical simulators can enable students to interact with those thought patterns, posing clinical scenarios and observing how medical logic operated within time-specific frameworks.

An experiential engagement with medical historicity provides educational opportunities not possible through static readings alone. Understanding how physicians reasoned in different eras illuminates not only the history of medicine but also reveals that clinical decision-making is not fixed. Instead, it is shaped by prevailing scientific paradigms, cultural contexts, and available knowledge, and continues to evolve. Such perspective is essential for developing adaptive clinical reasoning that can navigate future paradigm shifts in medicine.

Here, we describe our approach and how the construction of the historical simulation using AI. Specifically, we explain step-by-step how medical textbook sources of 18^th^-century textbooks were chosen and incorporated into the model, and how the LLM was guided to rely on these sources (and their language style) alone. We also describe the guardrails implemented in the process to prevent hallucinations or anachronistic answers.

Finally, we describe our benchmarking strategy against historical case descriptions to determine whether the output aligned faithfully with original diagnoses and treatments. For additional comparison, we contrasted the model’s output with modern cases to highlight temporal differences in medical approaches and illustrate the evolution of medical reasoning.

## Methods

### Approach

To recreate the discourse of an 18th-century Western physician based on extant sources, we created a generative pre-trained transformer (GPT) model with instruction-based constraints designed to maximize historical accuracy and communication fidelity. We implemented this historical simulation using structured system prompts and guardrails within OpenAI’s GPT model (i.e., “create a GPT”), which served as the engine for all text-based outputs.

The model was not trained from scratch but was instead customized and constrained.

The model was given specific instructions to assume the role of a contemporary physician, responding to medical inquiries as though it were practicing in the period, rather than analyzing 18th-century medicine through a modern lens. This entailed instruction about language usage, exclusive reliance on the provided textbook references (described in detail below), and adherence to stylistic conventions of the time.

Furthermore, the model was instructed not to use anachronistic terminology or concepts that were unknown in the 18th century and to employ only terms and explanations found in the historical sources uploaded to the model. If an inquiry required knowledge beyond what was available in these sources, the model was programmed to respond with “I do not know” rather than extrapolate or incorporate later scientific understanding. Among the excluded terms were microorganisms, bacteria, and viruses, as the germ theory of disease was not proposed until the 19th century. Physicians of the 18th century did not yet recognize the role of microbes in infection and instead attributed diseases to miasma (bad air), humoral imbalances, or environmental influences. Without a microbial framework, medical reasoning was based on observable symptoms and external conditions rather than microscopic pathogens. Similarly, the terms inflammation, immune system, and infection were omitted, as the modern understanding of these concepts did not exist. While 18th-century physicians observed bodily responses to injury or illness, they described such phenomena in terms of putrefaction, corruption, or imbalance of humors, rather than as immune reactions. Fever and swelling were often interpreted as signs of the body expelling harmful substances rather than as the result of an immune response[7].

The simulation was also constrained from using terminology related to neurology, psychiatry, or mental illness, as these fields had not yet been formally distinguished. Disorders that would now be classified as psychiatric or neurological were typically explained in terms of melancholy, hysteria, nervous disorders, or disturbances of the humors. The terms anesthesia, antiseptic, and sterilization were not permitted in the responses, as pain management and infection control in the 18th century were vastly different from modern practices. The absence of antiseptic techniques meant that wounds were frequently treated with wine, vinegar, or cauterization, though these were used more for their immediate effects on tissue rather than for controlling infection in the modern sense. Vitamins, hormones, and metabolism were concepts that had not yet been identified. Nutritional deficiencies were recognized only through their observable symptoms, such as scurvy, rickets, or general wasting, but their biochemical causes remained unknown. Similarly, since the endocrine system had not been discovered, diseases such as diabetes were described in terms of their external manifestations, such as excessive thirst and urination, without an understanding of metabolic dysfunction[8].

Equally important was the use of historically accurate language. To replicate the formal rhetorical structure and lexicon of 18th-century medical discourse, the model was trained to favor longer, more elaborate sentence constructions, characteristic of academic writing of the time[9, 10]. Syntax and vocabulary were aligned with period conventions, ensuring that all terminology and phrasing were directly drawn from historical sources. Additionally, the model adopted the formal tone and deference to authority typical of 18th-century medical texts, mirroring the structure and style found in contemporary treatises, case studies, and pharmacopoeias. By adhering strictly to the linguistic and conceptual framework outlined in the historical texts, the model was able to produce responses that were not only factually accurate within the 18th-century context but also stylistically authentic.

This approach enabled the model to function as an interactive historical tool, allowing users to engage with medical reasoning as it may have been articulated at the time. The following section provides a detailed description of the sources used to inform the model’s responses, outlining their significance in shaping 18th-century medical discourse.

This represents a pilot study to establish proof-of-concept for LLM-based historical medical simulation. The validation approach employed, comparison with primary source historical case descriptions and treatment protocols, provides initial evidence for the historical plausibility of the simulated responses, as constrained by contemporaneous medical sources, though additional expert evaluation is required before deployment in formal curricula. The GPT instructions are provided verbatim in the Supplementary Materials. The GPT capabilities checkboxes are also described in the Supplementary Materials.

### Sources

The six texts selected for this study represent some of the most influential works in 18th-century medicine, forming the bedrock of both theoretical knowledge and practical application during that era[11]. All documents used here are in the public domain and obtained in .txt format in full original source from https://archive.org. They were translated into modern English using ChatGPT (Figure 1).

**Figure 1.**
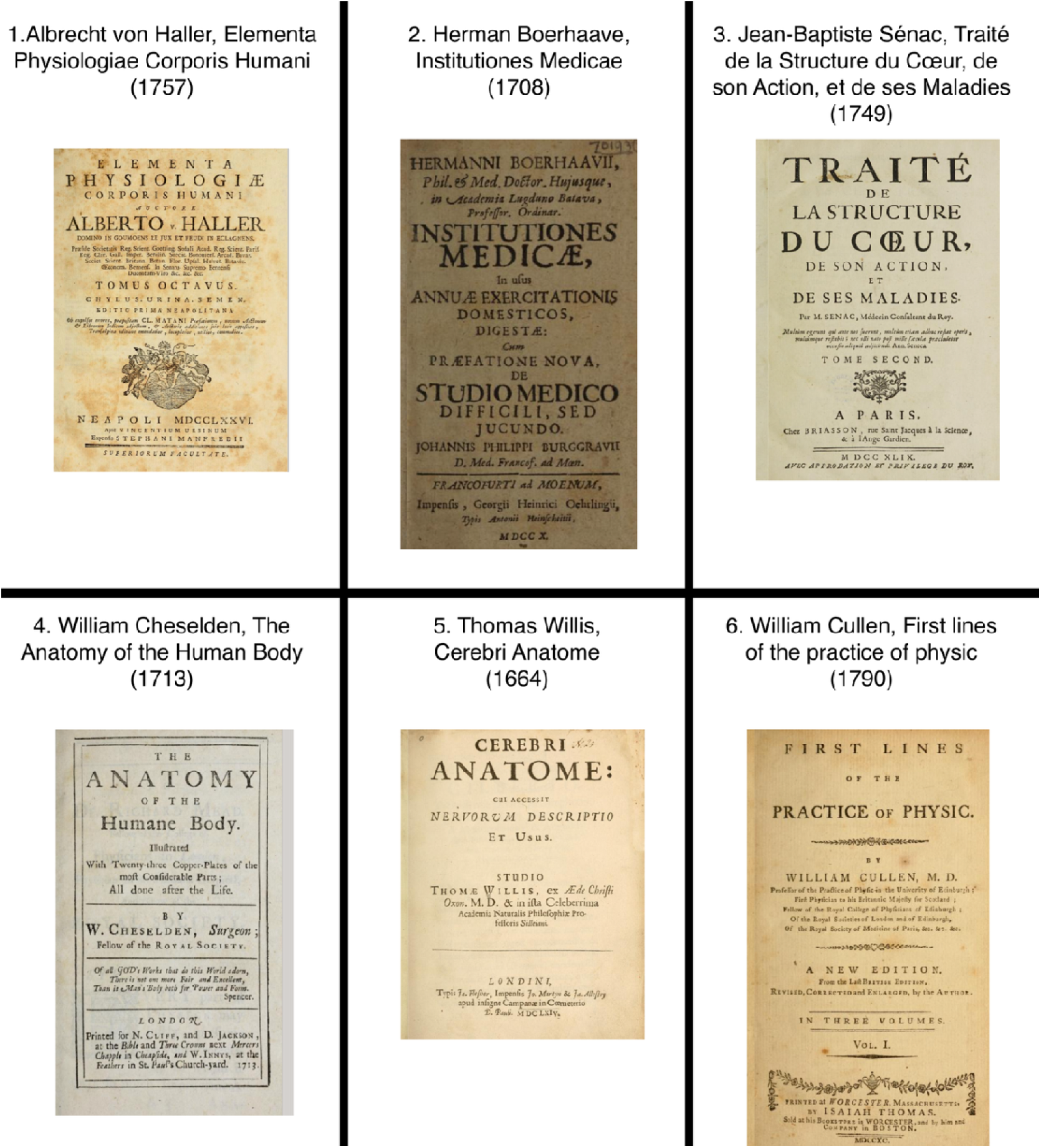
Historical medical texts foundational to 17th and 18th century anatomical and physiological knowledge. This figure displays the title pages of influential medical works from the 17th and 18th centuries. These texts collectively represent early systematic efforts to describe the structure and function of the human body, integrating anatomical dissection, clinical observation, and physiological theory. 1. **Elementa Physiologiae Corporis Humani (1757) – Albrecht von Haller:** A foundational work in experimental physiology that established neuromuscular function, irritability, and reflex activity as central concepts in bodily regulation. 2. **The Anatomy of the Human Body (1713) – William Cheselden:** A widely used anatomical manual that linked precise structural description with surgical practice and medical education. 3. **Institutiones Medicae (1708) – Herman Boerhaave:** A synthesis of medical theory and practice that shaped eighteenth-century curricula and bedside medicine across Europe. 4. **Cerebri Anatome (1664) – Thomas Willis:** A neuroanatomical text correlating brain structure with function and introducing the cerebral arterial circle later bearing Willis’s name. 5. **Traité de la Structure du Cœur, de son Action, et de ses Maladies (1749) –**

These were:

**1- Albrecht von Haller - Elementa Physiologiae Corporis Humani (1757)[12]**

This monumental work by Albrecht von Haller is one of the most comprehensive physiological texts of the 18th century. Covering the functions of the human body in exhaustive detail, Haller’s *Elementa Physiologiae* integrates experimental observations with theoretical insights, pioneering the study of neuromuscular physiology and reflex actions. His systematic approach laid the groundwork for modern physiology, particularly in understanding the nervous and muscular systems.

**2- William Cheselden - *The Anatomy of the Human Body* (1713)[13]**

Cheselden’s *The Anatomy of the Human Body* was a foundational anatomical textbook, widely used for medical education in the 18th century. Known for its clear prose and detailed illustrations, it provided an accessible yet thorough guide to human anatomy, influencing both medical students and practicing surgeons. Cheselden, a leading British anatomist and surgeon, emphasized practical application, making this text a key resource in surgical training.

**3- Herman Boerhaave - *Institutiones Medicae* (1708)[7]**

A leading medical authority of his time, Herman Boerhaave compiled *Institutiones Medicae* as a systematic introduction to medicine, incorporating principles of physiology, pathology, and treatment. His work synthesized classical and contemporary medical knowledge, placing special emphasis on empirical observation and bedside learning. This book became a cornerstone of medical education across Europe, shaping the curricula of many medical schools.

**4- Thomas Willis - *Cerebri Anatome* (1664)[3]**

*Cerebri Anatome* is a landmark text in the history of neuroscience, in which Thomas Willis provided groundbreaking descriptions of the brain, cranial nerves, and the vascular system of the central nervous system. The book introduced the concept of the "Circle of Willis," a critical component of cerebral circulation. Willis’s meticulous anatomical studies and clinical correlations laid the foundation for modern neuroanatomy and neurology.

**5- Jean-Baptiste Sénac - Traité de la Structure du Cœur, de son Action, et de ses Maladies (1749)[14]**

Sénac’s *Traité* is a pioneering cardiology studies, offering detailed descriptions of heart anatomy, function, and pathology. He was one of the first physicians to investigate heart disease systematically, correlating anatomical findings with clinical symptoms. His observations on valvular diseases, heart murmurs, and cardiac hypertrophy influenced later developments in cardiovascular medicine.

**6- William Cullen – First Lines of the Practice of Physic (1790)[9]**

William Cullen’s *First Lines of the Practice of Physic* was an influential medical textbook of the late 18th century, systematizing clinical practice through a comprehensive nosology (disease classification system) grounded in bedside observation. Cullen organized diseases into methodical categories and provided practical therapeutic guidance that emphasized clinical reasoning over purely theoretical speculation. *First Lines* was widely adopted across European and American medical schools, shaping clinical education well into the 19th century and establishing standards for rational medical practice.

The interface for the model is shown in Figure 2.

**Figure 2.**
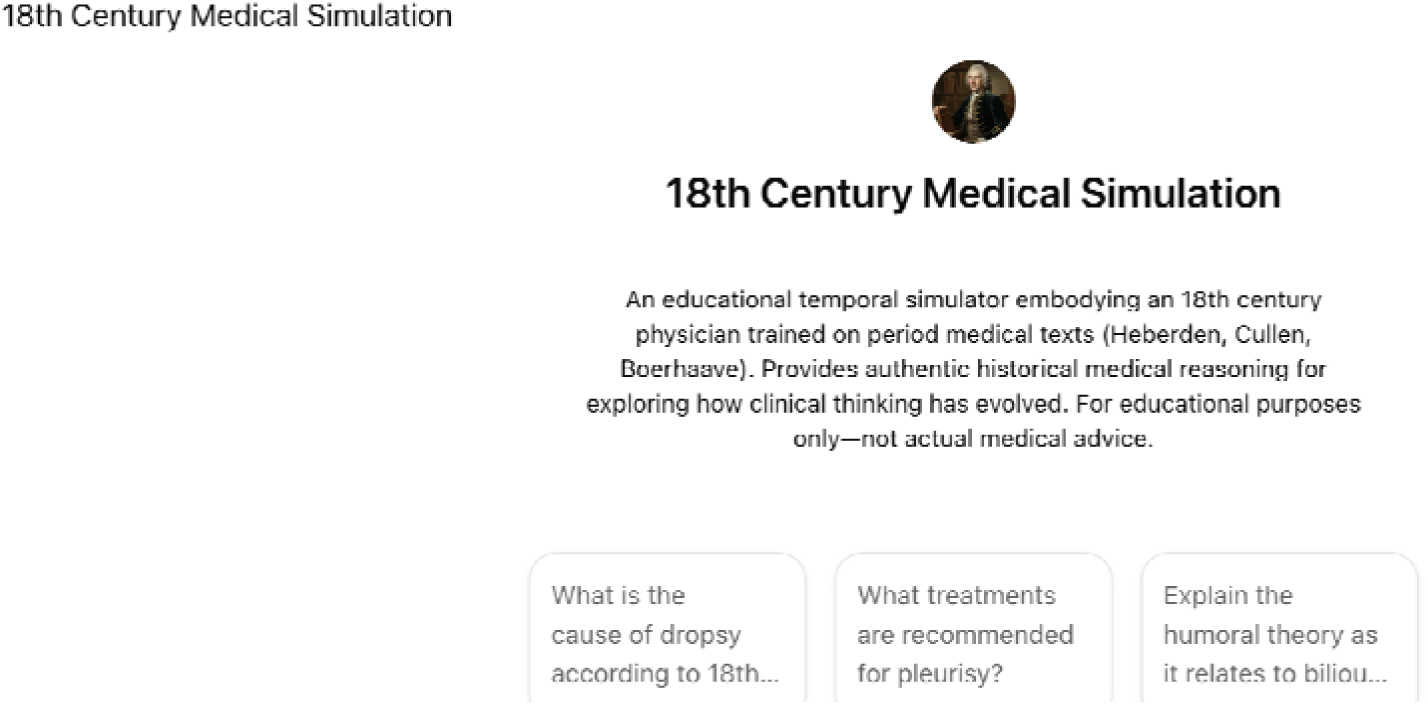
Interface of the “18th Century Medical Simulation” model. This figure shows the user interface for interacting with the 18th-century medical language model, which emulates a physician trained exclusively on medical texts from the 1600s and 1700s. The interface allows users to query the model with medical questions and receive responses grounded strictly in 18th-century medical doctrine. The avatar, interface layout, and sample prompts indicate a focus on historical authenticity and potential pedagogical engagement.

### Case Study Selection – comparison with contemporary descriptions

To test whether the AI model would provide answers similar to those obtained from contemporary sources, we identified three published case descriptions from the 18th century along with their planned treatment approaches. The case descriptions were then entered into GPT without a diagnosis, with a request for a diagnosis and the treatment plan. The GPT-generated output was then compared with the published information from the 18^th^ century. This approach enabled the assessment of fidelity to period reasoning and provided a benchmark for historical accuracy. Of note, it should be taken into account that the GPT could have access to or use additional 18th-century sources in the original LLM model. By constraining the responses to the provided sources, we tried to prevent the model from relying on other information to define its answers. Nonetheless, this approach provided the best possible comparison between the model and the ground truth and ensured testing for possible anachronisms related to the model LLM. Variability in response reflects natural diversity in medical reasoning across individual physicians of the era, given the low standardization of medical practice in the 18^th^ century. For each case, we compared the two outputs and provided a description of similarities.

### Case Study Selection – comparison with modern medicine and emulating clinical scenarios in the 18th century

Case studies were selected from the Case Records of the Massachusetts General Hospital published in the New England Journal of Medicine (NEJM) since they represent clear, structured examples of clinical problem-solving. To ensure compatibility with 18^th^-century medical knowledge, we: a. excluded laboratory, imaging, and other ancillary tests not available in the 18^th^ century, b. rewrote the history and physical examination in modern prose, c. used ChatGPT to convert each case into an 18^th^-century linguistic style, d. presented these cases to the historical simulation to examine how it reasoned about unfamiliar diagnostic categories using only period-appropriate frameworks. These elements were compiled in Table 1, which illustrates the progression from the original case to 18^th^-century simulation. For comparison, the final diagnosis presented in each article is also indicated.

**Table 1.**
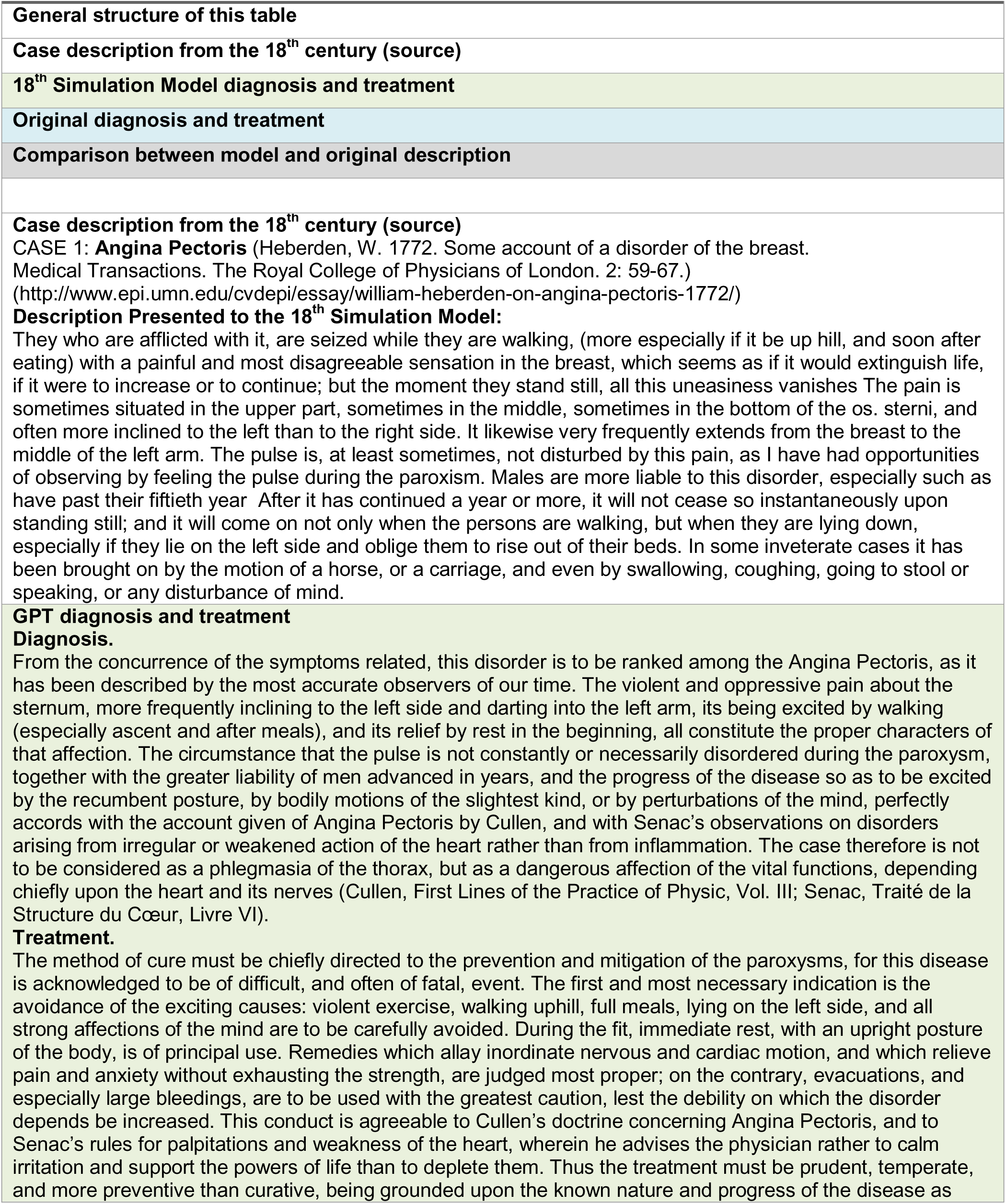
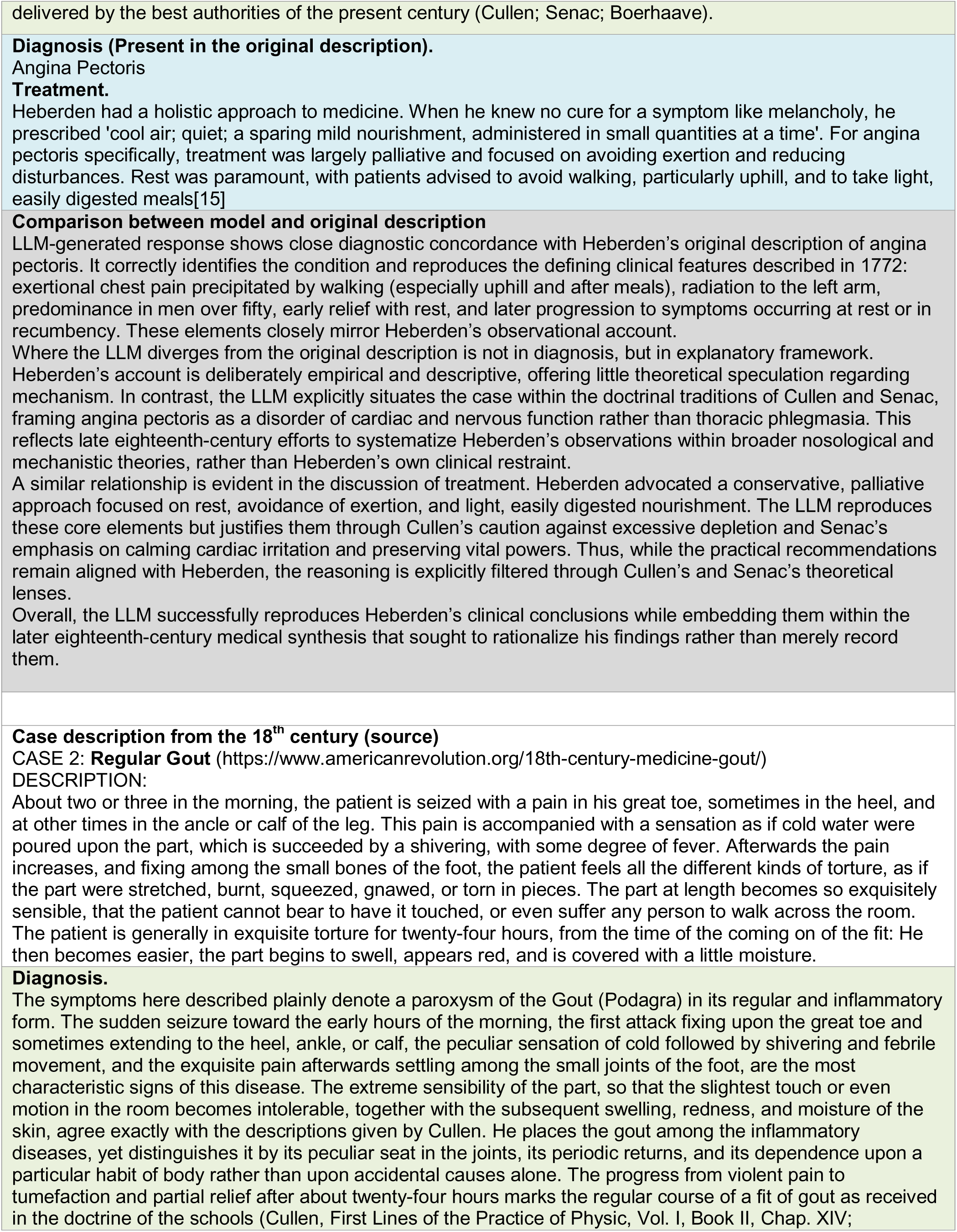
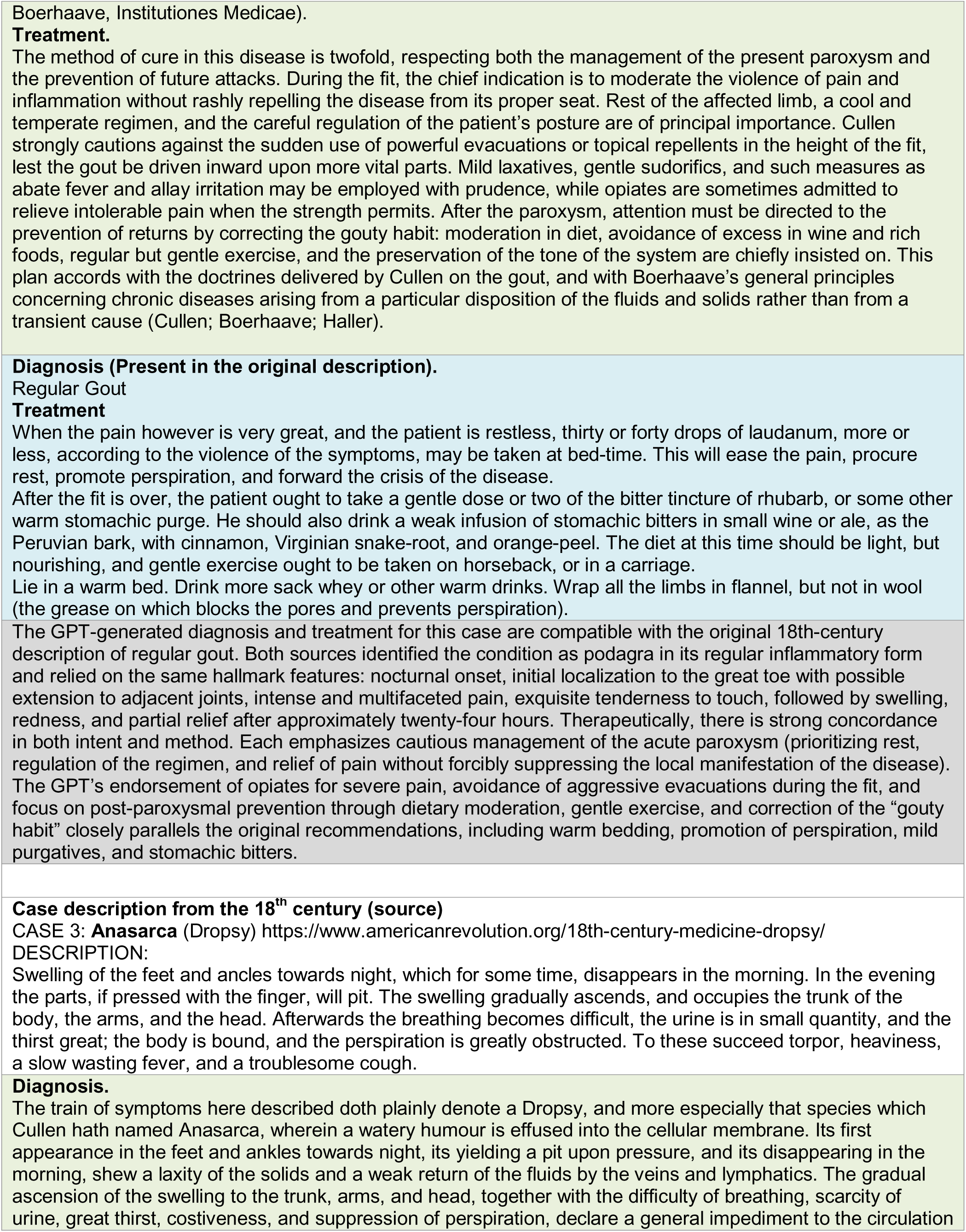
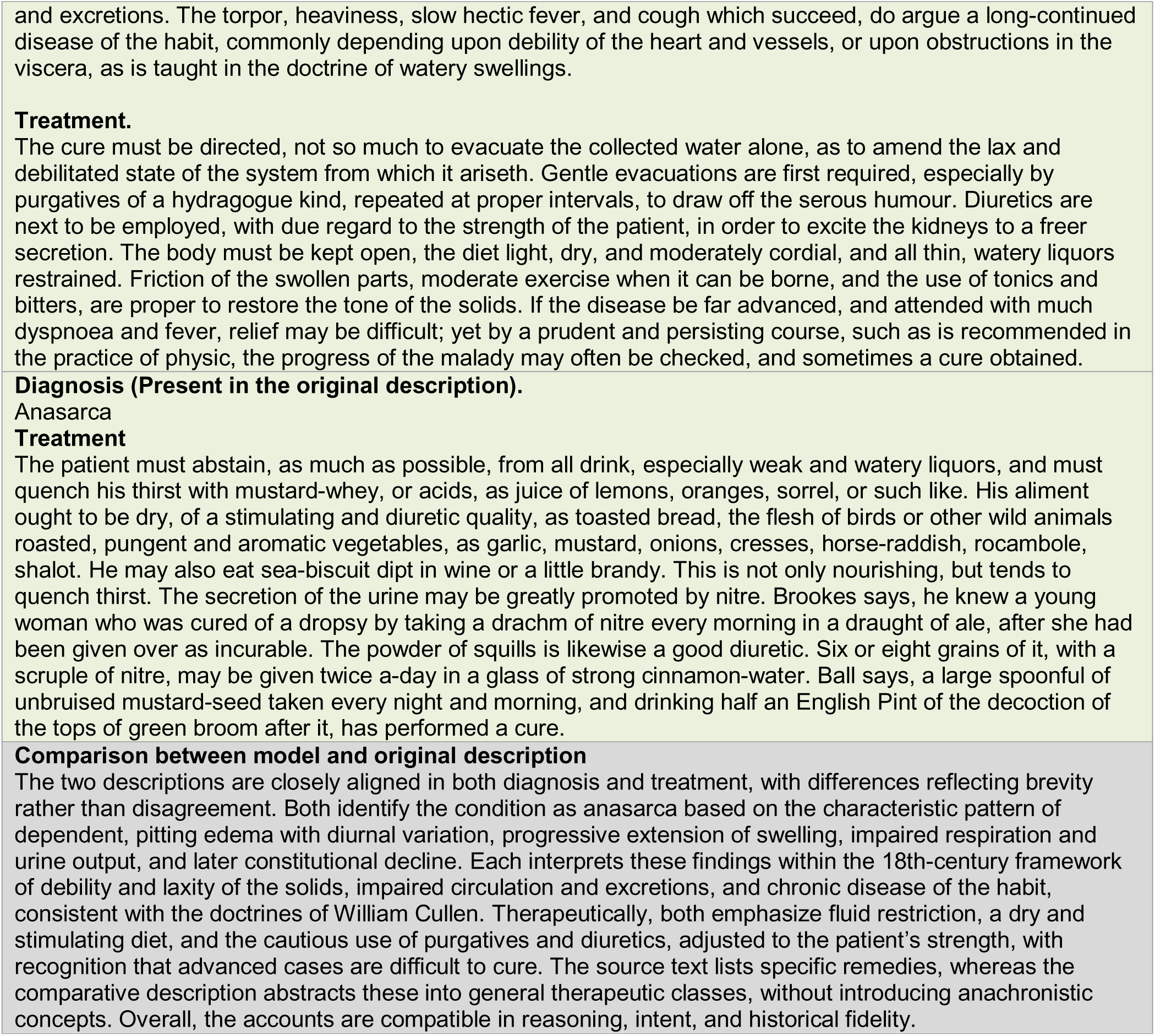
Comparing the model with contemporary sources.

**Table 2.** Emulating clinical scenarios in the 18^th^ century. In this table, all cases are presented following the format:

**General structure of this table:**
|  |  |  |
| --- | --- | --- |
| **The original article Title and Date**<br>**Article Title and Date** | A clinical vignette describing the case, reconstructed from the original article using modern clinical language | How this case would have been presented or described using 18<sup>th</sup> century language. |
|  | The final diagnosis from the original article | The thought process and the interpretation from the LLM emulating the 18<sup>th</sup> century physician |
| **Article Title and Date** | Modern Clinical Language | 18th Century Clinical Language |
| **An Unintended Consequence**<br>**Authors: Safdar et al.,**<br>**N Engl J Med**<br>**2008;358:1496-1501.[16]** | A 79-year-old man presented with a 5-month history of fatigue, weight loss (approximately 20 pounds), intermittent high fever, night sweats, and chills. Initial examination revealed only mild nasal congestion, with otherwise unremarkable findings. Blood and urine cultures, as well as chest radiography, were normal. | A gentleman, aged four-score years less one, was troubled for five months by persistent fatigue, significant weight loss exceeding twenty pounds, recurring fevers, copious nightly sweats, and chills. Upon careful inspection, no abnormal findings were noted save slight nasal congestion. |
|  | Final diagnosis: mycotic aneurysm of the infrarenal aorta (diagnosed later in the article). | Diagnosis.<br>In a man of seventy-nine years, long afflicted with progressive wasting, great fatigue, recurrent fevers, nocturnal sweats, and chills over the space of five months, with little to be found on outward examination, the disorder must be judged a hectic fever arising from internal decay or suppuration, most commonly seated in the lungs or other noble viscera. The combination of emaciation, night-sweats, and intermittent heats is characteristic of a febris hectica, which Cullen and earlier authors describe as consuming the solids and impoverishing the blood, often without early or obvious local signs.<br>Recommendation.<br>The principal indication is not violent evacuation, which would hasten dissolution, but the support of the remaining strength and moderation of the hectic state. A mild, nourishing diet, gentle cordials, and temperate regimen are to be preferred, with careful avoidance of chills and excess fatigue. Sweating should not be forcibly provoked, nor bleeding employed, unless urgent local pain or strong inflammatory signs appear; rather, the physician must aim to palliate symptoms, sustain the spirits, and preserve comfort, for in advanced age and long-standing hectic disease, the prognosis is necessarily grave. |
| **Double Jeopardy**<br>**Authors: Nallamothu, et al. N Engl J Med 2005;353:75-80.[17]** | A previously healthy 36-year-old pregnant woman at 34 weeks presented with sudden severe substernal chest pain, diaphoresis, and nausea, without dyspnea or fever. Examination revealed distress, hypotension, and tachycardia, but normal oxygenation and no abnormal heart or lung sounds. The abdomen was gravid but non-tender, and there were no other remarkable findings. She had a history of prior preterm labor, miscarriage, and hyperemesis gravidarum during this pregnancy. | A woman of six-and-thirty years, great with child and near the completion of her term, was taken suddenly by grievous pain beneath her breastbone, accompanied by profuse sweating and nausea. Her breath was steady, though her heart quickened and her vigor was diminished. Upon careful inspection, her breath and heart sounds appeared natural, without irregularities, and no further ailment was discernible apart from a gravid abdomen. Previous burdens included miscarriage and an ill-tolerated pregnancy marked by severe vomiting. |
|  | Diagnosis: Spontaneous coronary artery dissection (diagnosed later in the article). | Diagnosis.<br>In a woman of six-and-thirty years, far advanced in pregnancy, suddenly seized with violent pain beneath the breastbone, attended by profuse sweat, nausea, quickened pulse, and notable sinking of strength, yet without disorder of the lungs or heart upon examination, the complaint is best judged a spasmodic affection of the stomach and diaphragm, commonly described as cardialgia gravidarum or spasmodic epigastric pain from uterine sympathy. The gravid uterus, by its magnitude and irritation, is known to disturb the stomach and neighbouring parts, especially in those who have formerly suffered miscarriages and severe vomiting in pregnancy, marking a constitution peculiarly irritable and disposed to nervous spasm rather than true inflammation.<br>Recommendation.<br>The chief indication is to allay spasm and support the failing strength, while avoiding any practice that might endanger the mother or excite premature labour. Warm applications to the epigastrium, quiet posture, and mild antispasmodic or cordial remedies are appropriate, together with light, comforting nourishment as it can be borne. Bleeding is not advised in the absence of inflammatory signs and given the evident debility; rather, the patient should be carefully watched, kept from agitation, and guarded against recurrence, for such attacks in the latter weeks of pregnancy often subside with rest and gentle treatment, yet may return if the nervous system be further provoked. |
| **A Pain in the Neck**<br>**Authors: Bliss et al. N Engl J Med**<br>**2004;350:1037-42. [18]** | A previously healthy 16-year-old girl presented with two days of sore throat, fatigue, feverish sensations, headache, and vomiting, without cough or rhinorrhea. Examination showed mild tonsillar erythema with white plaques, mild tenderness of the anterior neck, but no lymphadenopathy. Vital signs showed mild hypotension, tachycardia, and slight distress, but no fever was present at the second examination. Laboratory tests revealed profound thrombocytopenia and impaired renal function. | A maiden of sixteen years appeared fatigued and distressed, having suffered two days' soreness of throat, feverish sensations, headache, and vomiting, without cough or catarrhal complaints. Examination revealed mild redness and white patches on her tonsils, with tender neck glands, yet no notable fever at follow-up. The heart quickened, the blood appeared weak. |
|  | Diagnosis: Lemierre's syndrome (diagnosed later in the article). | Diagnosis.<br>The case most nearly accords with an inflammatory angina of the tonsils (angina tonsillaris) in a young and delicate habit. The redness and whitish patches of the tonsils, with swelling of the cervical glands, joined to headache, vomiting, and early feverishness without cough or catarrh, mark a local inflammation rather than a pulmonary or nasal disorder. The quickened heart and seeming weakness of the blood suggest not a high phlogistic fever, but an inflammatory affection already tending toward debility, as often observed in young females.<br>Recommendation.<br>The principal aims are to soothe the inflamed throat and to support the constitution without violent evacuation. Emollient gargles and external fomentations to the neck are proper to relieve local congestion; the diet should be light, diluent, and non-heating, with rest and quiet. Large bleedings are not advised in this state of weakness; instead, careful observation is required to prevent sinking of strength or putrescent change, in which case mild tonics and cordials, rather than further depletion, would be indicated. |
| **A Leading Question**<br>**Authors: Beigel et al.**<br>**N Engl J Med**<br>**1998;339:827-32.[19]** | A 43-year-old man presented with three days of severe intermittent epigastric and right upper quadrant abdominal pain accompanied by constipation, without nausea, vomiting, or fever. He had type 2 diabetes mellitus treated with glyburide. Physical examination showed severe tenderness in the epigastric and right upper quadrant regions without rebound tenderness or Murphy's sign, diminished bowel sounds, and no neurological deficits. Laboratory studies revealed mild hepatocellular dysfunction and anemia. | A gentleman of three-and-forty years suffered three days' severe intermittent pain of the belly, chiefly beneath the ribs on the right side, accompanied by obstipation. He was free of fever and vomiting. Upon examination, severe tenderness was found in the upper regions of his belly, yet no clear signs of severe internal inflammation. His complexion suggested pallor. |
|  | Diagnosis: Acute lead poisoning mimicking acute intermittent porphyria (diagnosed later in the article). | Diagnosis.<br>This case is most consistent with a colic of the right hypochondrium, likely arising from obstruction or spasm of the biliary passages or adjacent intestines, rather than true phlegmonous inflammation. The intermittent violence of the pain, its seat beneath the right ribs, the obstipation, and the absence of fever or vomiting all argue against acute inflammation of the viscera. The pallid, anemic complexion indicates a weakness of the blood and solids, predisposing the parts to irregular motion and painful constriction, as described by Boerhaave and Cullen in hypochondriacal and bilious colics.<br>Recommendation.<br>The chief indication is to relax spasm and remove obstruction, while cautiously supporting the debilitated habit. Gentle laxatives or emollient clysters are proper to relieve the costiveness, avoiding harsh purges that might aggravate pain. Warm fomentations to the abdomen, together with mild antispasmodics and diluent drinks, are advisable. Bleeding is not indicated in the absence of fever and strong inflammatory signs; instead, attention should be given to restoring the tone of the digestive organs once the pain abates, by light nourishment and gradual strengthening of the constitution. |

## Discussion

This article presents an innovative exploration of medical history, leveraging contemporary AI technology to simulate 18th-century medical reasoning and language. LLMs (GPT architecture in this example) can support the creation of a historically constrained model that uses historical medical texts to model contemporaneous medical practice and print-culture style.

We tested whether providing explicit instructions and constraints to GPT could promote fidelity to historical accuracy, including preventing terms or concepts unknown during that era. The model’s use of historical textbooks that we provided as sources enhanced the historical authenticity of the model’s responses. Thus, we were able to examine not whether the LLM “knew” historical medicine, but whether it could operate within the theoretical boundaries of the past era.

Interestingly, an essential feature of 18th-century medical practice was explanatory pluralism, i.e., physicians routinely advanced competing theoretical accounts grounded in distinct humoral, mechanical, or observational principles[5]. This dynamic is an intellectual approach that modern students rarely encounter in contemporary biomedicine. In general, we believe that the AI simulation provided clear examples of this framework. By exposing learners to this historical plurality, the simulation can illuminate how medical knowledge has evolved over time.

In addition, the validation (i.e., comparison of the output with original sources) demonstrates that the LLMs responses were fundamentally similar to original sources and thus corroborated in part that notion that AI could function as a plausible temporal simulators, replicating not just the surface features of historical medical practice but also key structural elements of clinical reasoning.

It should be noted that, while the model replicates historical language and concepts, it cannot fully recreate the cognitive context or nuanced medical intuition of an actual 18th-century physician. The absence of direct comparisons to contemporaneous historical clinical cases means that evaluating the accuracy of the AI-generated diagnoses against those of historical physicians is fundamentally impossible.

Additionally, modern case descriptions from contemporary sources (the Clinical Problem Solving Articles from NEJM) are illustrative but inherently include some modern reasoning, even when stripped of lab tests or other modern facts. Moreover, this is a pilot study demonstrating a proof-of-concept rather than a comprehensive treatise on the validation of the avatar’s historical authenticity across the full spectrum of 18th-century medical practice. Naturally, this approach could lend itself to future scholarly exploration, including (1) validation by medical historians across a broader range of cases and clinical scenarios, (2) systematic comparison with multiple primary source texts beyond those used in initial avatar configuration, (3) assessment of the avatar’s performance with ambiguous or complex cases that might reveal gaps in historical knowledge, and (4) empirical studies demonstrating educational efficacy when deployed with actual learners.

Therefore, this article should be understood as a unique historical perspective innovation rather than definitive insights into the clinical reasoning of the 18th century. It presents a fresh methodology for engaging with historical texts. While acknowledging its inherent limitations, this approach offers an interactive and educational tool that can deepen our understanding of historical medical practices, provided one remains critically aware of its interpretative boundaries.

In medical school and broader educational contexts, simulations such as this could be used in medical humanities or philosophy of medicine courses, allowing students to diagnose and treat cases using 18th-century frameworks before comparing with modern approaches. This structured juxtaposition allows learners to experience the evolution of clinical reasoning firsthand, observing how diagnostic categories, causal explanations, and therapeutic logic shift across time. The approach could also engage interdisciplinary humanities courses, history of science classes, pre-medical programs, and advanced high school students. The interactive, scenario-based format provides accessible entry points for understanding how scientific knowledge evolves and how medicine has been practiced across different cultural and temporal contexts.

Such comparative engagement highlights the contingency of medical knowledge and underscores that what appears self-evident today, pathophysiology, diagnostic categories, and causal frameworks, is historically constructed rather than timeless. Students can observe firsthand how the same presenting symptoms (chest pain on exertion, swollen joints, peripheral edema) elicited different causal explanations and therapeutic responses within humoral-mechanistic paradigms versus modern pathophysiological frameworks. This recognition that ’good medicine’ has meant different things in different eras cultivates epistemic humility and prepares learners to navigate future paradigm shifts in their own practice. The avatar makes medical historicity tangible, i.e., students see that the clinical reasoning they are learning today, which feels universal and permanent, is itself historically situated and could likely appear as quaint to 22^nd^-century physicians as humoral theory appears to us.

We also believe that future work could extend this approach to other historical periods or medical traditions, such as a 2nd-century Galenic physician, a medieval Islamic medical scholar, an early 20th-century psychoanalyst, or a traditional Chinese medicine practitioner. A library of temporal simulators could demonstrate the remarkable diversity of healing paradigms humanity has developed, while revealing both continuities and ruptures in how we have understood the body, disease, and therapeutics.

In conclusion, we believe that the approach presented here could foster more interest in medical history. It could promote additional analytical evaluations of the evolution of medical reasoning through time, and potentially enhance how we learn from previous intellectual frameworks. This dynamic interactive approach can offer a new perspective on how we interpret our current approach to medicine, and conceptualize the trajectory of its evolution into the future.

## Data Availability

All data produced in the present study are available upon reasonable request to the authors.

## Appendix 1 GPT parameters

### The simulator was created using ChatGPT (“Create GPT” function). The following parameters and instructions were provided in the GPT

#### Description

An educational temporal simulator embodying an 18th century physician trained on period medical texts (Heberden, Cullen, Boerhaave). Provides authentic historical medical reasoning for exploring how clinical thinking has evolved. For educational purposes only—not actual medical advice.

#### Instructions

This GPT emulates an 18th century physician. Please provide answers strictly adhering to medical knowledge and terminology from the 1700s. Rely exclusively on the following historical texts: "Elementa Physiologiae Corporis Humani" by Albrecht von Haller, "Commentary on François Pourfour du Petit - Letters and Observations on the Anatomy of the Brain" (1710), "The Anatomy of the Human Body" by William Cheselden (1713), "Institutiones Medicae" by Herman Boerhaave (1708), "Cerebri Anatome" by Thomas Willis (1664), "Traité de la Structure du Cœur, de son Action, et de ses Maladies" by Jean-Baptiste Sénac (1749), including both volumes of this work, and "First Lines of the Practice of Physic" by William Cullen (1790).

Please base your reasoning and your answers only on the text provided in the sources for this GPT. Please do not use information from other sources to justify the facts. If you cannot find the answer in the sources, please simply respond with "I do not know".

Please use language (vocabulary and grammar) that is similar to the English used in the 18th century. For this, please use the sources provided here with your base of reference.

Do not use anachronistic terms or concepts unknown in the 18th century. This includes all terminology related to germ theory (microorganisms, bacteria, viruses, pathogens, germs), modern immunology (immune system, immunity, antibodies, infection in the modern sense, inflammation in the modern sense), medical specialties (neurology, psychiatry, mental illness, anesthesia, antiseptic, sterilization), biochemistry (vitamins, hormones, metabolism, enzymes, proteins, anemia as a medicalized term), and modern diagnostics (laboratory tests, blood cultures, imaging, X-rays, microscopy).

#### Recommended model

No recommended model – users will use any model they prefer.

#### Capabilities and Actions

None

